# Vaccine-induced immune thrombotic thrombocytopenia (VITT) is mediated by a stereotyped clonotypic antibody

**DOI:** 10.1101/2022.03.28.22272975

**Authors:** Jing Jing Wang, Bridie Armour, Tim Chataway, Alexander Troelnikov, Alex Colella, Olivia Yacoub, Simon Hockley, Chee Wee Tan, Tom Paul Gordon

## Abstract

Vaccine-induced immune thrombotic thrombocytopenia (VITT) is a rare thromboembolic complication of adenoviral-vectored SARS-CoV2 vaccines, mediated by antibodies directed against platelet factor 4 (PF4). Given their causal role in VITT, identification of the molecular composition of anti-PF4 antibodies is crucial for developing better diagnostics and treatments. Here, we utilised a novel proteomic workflow to analyse the immunoglobulin variable (IgV) region composition of anti-PF4 antibodies at the level of the secreted proteome. Serum anti-PF4 IgG antibodies from five patients with VITT triggered by ChAdOx1 nCoV-19 vaccination were affinity purified by PF4-coupled magnetic beads and sequenced by mass spectrometry. We revealed a single IgG heavy (H)-chain species paired with a single lambda light (L)-chain species in all five unrelated patients. Remarkably, all L-chains were encoded by the identical IGLV3-21*02 gene subfamily with identical L-chain third complementarity determining region (LCDR3) lengths. Moreover, striking stereotypic features were also identified in heavy-chains anti-PF4 antibodies characterised by identical HCDR3 length and homologous sequences. In summary, we unravelled the molecular signature of highly stereotyped clonotypic anti-PF4 antibodies, indicating shared pathways of antibody production in VITT patients. These discoveries are critical to understand the molecular basis of this serious condition and develop novel therapies aimed at removing pathogenic clones.

**KEY POINTS:**

- Anti-PF4 antibodies in VITT comprise highly stereotyped clonotype
- A single IGLV3-21*02 encoded light chain is found in unrelated patients

## Introduction

The syndrome of vaccine-induced immune thrombotic thrombocytopenia (VITT) is a rare thromboembolic complication of adenoviral-vectored SARS-CoV2 vaccines ChAdOx1 nCoV-19 (AstraZeneca) and Ad26.COV2.S (Janssen/Johnson & Johnson) mediated by antibodies directed against platelet factor 4 (PF4).^1-5^ The mechanisms by which the adenoviral DNA vectors break immune tolerance to PF4 and trigger B-cell clonal expansion and secretion of anti-PF4 IgGs are under intense investigation and likely involve formation of immunogenic complexes of PF4 with vaccine components in a pro-inflammatory setting.^6^ Pathogenic anti-PF4 IgGs subsequently form circulating immune complexes with PF4 tetramers which are thought to drive thrombotic events by Fc gamma receptor IIa (FcγRIIa)-dependent platelet activation and to activate granulocytes to release procoagulant neutrophil extracellular traps (NETS).^6-8^ Serum anti-PF4 antibodies are mostly transient and appear in serum within days of vaccination, suggesting a recall immune response on memory B-cells.^9^

Given their causal role in VITT, identification of the molecular composition of the anti-PF4 antibodies and their antigenic target(s) is crucial for developing better diagnostics and treatments and for precise tracking of PF4-specific B-cell clones and secreted clonotypes. In a key advance, Huynh et al, have mapped the antibody-binding site to a single conformational epitope on the PF4 molecule which is located within the heparin-binding site and distinct from epitopes bound by serum from patients with heparin-induced thrombocytopenia (HIT).^10^ Moreover, a recent intact mass spectrometric analysis of anti-PF4 IgGs in patients with VITT and HIT revealed expression of monoclonal and oligoclonal light chains in the former as distinct from a polyclonal light-chain pattern in the latter. While intact light-chain mass measurements were performed to inform clonality, direct amino acid (aa) sequencing of light-or heavy-chains was not investigated in this study.^11, 12^

We have developed a novel proteomic workflow in our facility based on high-resolution de novo mass spectrometric sequencing of immunopurified serum antibodies to identify their immunoglobulin variable (IgV) subfamily expression profiles; clonotypical light and heavy-chain third complementarity determining region (LCDR3 and HCDR3) aa sequences as barcodes for clonal tracking; and V-region aa replacement mutational signatures as molecular markers of antigen-driven intraclonal diversification.^13-16^ Here, we have utilised a proteomics discovery approach (Figure 1A) to profile serum anti-PF4 antibodies in VITT patients and unexpectedly reveal stereotypic (also termed public) LCDR3 and HCDR3 aa sequences with near perfect light chain stereotypy. This points to highly convergent pathways of anti-PF4 antibody production and potential translation of shared CDR3 peptide “barcodes” to novel molecular biomarkers for these highly pathogenic clonotypes.

**Figure 1.**
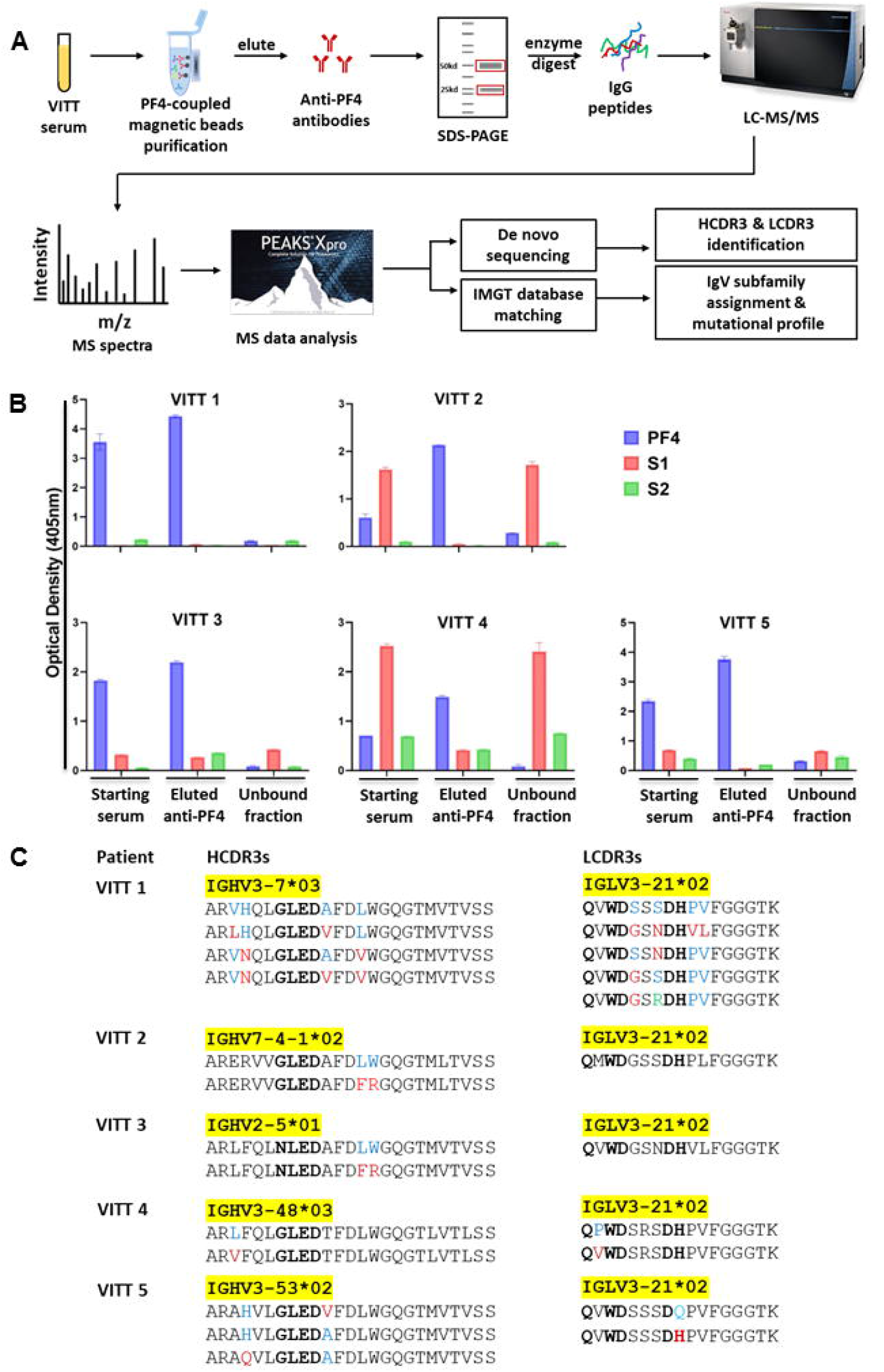
Mass spectrometry-based characterisation of PF4-specific clonotypic antibodies. (A) Proteomics workflow to identify molecular signatures of anti-PF4 antibodies. PF4-specific Igs are purified from serum of VITT patients using PF4-coupled magnetic beads. Heavy (H) and light (L)-chains are separated by reduced sodium dodecyl-sulfate polyacrylamide gel electrophoresis (SDS-PAGE) and excised and digested with enzymes to generate peptides for liquid chromatography mass spectrometry/mass spectrometry (LC-MS/MS). Ig variable region peptide sequences are analysed by combined de novo sequencing and IMGT database matching. (B) Specificity of purified anti-PF4 antibodies. Magnetic bead-purified anti-PF4 Igs from five individual VITT patient sera are tested by PF4, SARS-CoV-2 spike S1 and S2 ELISAs. Data are shown as mean ± SD (n=2). (C) Clonotypic H and L-chain third complementarity-determining region (CDR3) signatures. Ig variable region subfamilies of PF4-specific antibodies as highlighted in yellow are assigned by IMGT database matching. HCDR3 and LCDR3 amino acid sequences from five individual VITT patients are identified by de novo sequencing. Bold amino acids in HCDR3s and LCDR3s denote shared motifs across unrelated patients, and amino acids in color denote amino acid replacement mutations found in individual patient HCDR3 and LCDR3 regions.

## Study design

### Human subjects

Diagnostic serum specimens were obtained from five patients with VITT (mean age 65 years (range 49–88)) whose clinical and serological findings are summarised in Table 1. The Asserachrom HPIA IgG Stago and the Lifecodes PF4 IgG Immucor assays were used for diagnostic detection of anti-PF4 antibodies.^17^ All five cases satisfied the case definition criteria for “Definite VITT”, according to the UK Expert Haematology panel and Level 1 Certainty Determination for Thrombosis with Thrombocytopenia Syndrome (TTS), as outlined in the updated Brighton Collaboration Case Definition for TTS.^18, 19^ Four of the five cases were characterised by thrombosis in critical/unusual sites, mainly cerebral venous sinus thrombosis (CVST) and splanchnic vein thrombosis (SVT). All study subjects gave written informed consent prior to inclusion in the study. The study was approved by the Clinical Ethics Committee of the Flinders Medical Centre using de-identified diagnostic sera (approval number 39.034).

**Table 1:** Laboratory and clinical features of ChAdOx1 nCoV-19 (AstraZeneca)-associated VITT patients.

| **Patient code** | **Timing of symptom onset after vaccination (days)** | **Platelets (x10<sup>9</sup>/L) \*** | **Fibrinogen (g/L) §** | **D-dimer (mg/L) #** | **Thrombotic Features** | **Asserachrom HPIA IgG, Stago (OD) ¶** | **Lifecodes PF4 IgG, Immucor (OD) \*\*** |
| --- | --- | --- | --- | --- | --- | --- | --- |
| VITT 1 | 12 | 11 | 1.1 | 44.7 | CVST with secondary ICH | 2.43 | 3.14 |
| VITT 2 | 15 | 13 | 1.5 | >80 | CVST, PE, SVT | 0.73 | 3.39 |
| VITT 3 | 13 | 57 | 1.3 | 52.4 | Bilateral DVT | 2.24 | 0.89 |
| VITT 4 | 13 | 60 | 1.0 | >80 | SVT requiring bowel resection | 2.18 | 1.17 |
| VITT 5 | 12 | 40 | 1.4 | 35 | CVST, DVT | 1.46 | 1.08 |

VITT indicates vaccine-induced immune thrombotic thrombocytopenia; CVST, cerebral venous sinus thrombosis; DVT, deep vein thrombosis of legs; ICH, intracerebral haemorrhage; PE, pulmonary embolism; SVT, splanchnic vein thrombosis
\*Reference range: 150-450 x10<sup>9</sup>/L
§ Reference range: 1.5-4.0 g/L

### Proteomic profiling serum anti-PF4 antibodies

As denoted in the proteomics workflow (Figure 1A), anti-PF4 IgGs were immunopurified by PF4 (ChromaTec, Greifswald, Germany)-coupled magnetic beads (ThermoFisher) from serum of patients with VITT. Prior to sequencing, monospecificity of anti-PF4 IgGs was verified by testing starting serum (diluted 1/100), bead-purified anti-PF4 antibody fraction and unbound fractions using ELISAs coated with individual PF4, SARS-CoV-2 spike S1 and S2 proteins (the Native Antigen Company, UK) at 4 μg/ml., purified anti-PF4 IgGs were then separated by SDS-PAGE; heavy- and light-chain bands excised for in-gel digestion with trypsin and chymotrypsin, respectively; and analysis of peptides performed in a state-of-the-art Thermo Orbitrap Fusion Lumos Tribrid mass spectrometer with peptide de novo sequencing and IMGT database matching performed by Peaks Studio XPro as detailed previously ^15^ with modifications. Annotated de novo peptide spectra from HCDR3 and LCDR3 clonotypic peptide barcodes are shown in Supplementary Figure 1. For more details about the experimental and bioinformatic methods, see Supplementary Methods.

## Results and discussion

Prior to sequencing, monospecificity of anti-PF4 IgGs was evaluated by in-house ELISAs. There was no cross-reactivity between eluted, purified anti-PF4 IgGs and S1 and S2 proteins in the VITT serum samples, consistent with a recent report ^20^ (Figure 1B).

Mass spectrometric sequencing of anti-PF4 Igs revealed a single IgG H-chain species paired with a single lambda L-chain species in all five unrelated patients. Remarkably, all L-chains were encoded by the identical IGLV3-21*02 gene subfamily and showed identical LCDR3 peptide lengths consistent with a high degree of L-chain stereotypy. Notably, the shared IGLV3-21*02 allele expresses an acidic (negatively charged) DDxD motif in the CDR2 region which we suggest may be of potential importance in antibody binding to the positively charged PF4 epitope.^10^ Another shared aa motif of interest QxWD is located in the LCDR3 region (Figure 1C). The roles of these putative binding motifs await confirmation by formal structural studies. Interestingly, a stereotypic IGLV3-21 B-cell receptor defines a subgroup of chronic lymphocytic leukemia with a poor prognosis.^21^

HCDR3 peptides are specific clonotypic markers of serum antibodies. However, clonotypic HCDR3 peptides cannot be identified by conventional database matching due to the lack of reference databases in IMGT for rearranged variable (V)-diversity (D)-joining (J) segments. Accordingly, we have developed an advanced de novo sequencing workflow to identify HCDR3 clonal barcode peptides and identified striking stereotypic features characterised by identical HCDR3 lengths and homologous sequences, together with a shared binding motif G/NLED which was located in IGHD regions known to confer antigen binding specificity ^22^ (Figure 1C). Notwithstanding these convergent HCDR3 regions, individual patient anti-PF4 Ig proteomes were encoded by distinct IGHV subfamilies including 3-7, 7-4, 2-5, 3-48 and 3-53 (Figure 1C), emphasising the critical role of HCDR3s in PF4 epitope binding as opposed to the divergent IGHV regions.

Amino acid replacement mutations were also found in individual patient HCDR3 and LCDR3 regions (Figure 1C), consistent with a model of PF4 antigen-driven intraclonal diversification as observed for systemic autoantibodies in lupus and Sjögren’s syndrome.^13, 14, 23^ Additional glutamic acid (E) and aspartic acid (D) replacement mutations of potential binding significance were identified in the IgV regions of H-and L-chains, suggesting recall immune responses on PF4-specific memory B cells (data available upon request).

The finding of a stereotyped clonotypic anti-PF4 antibody in this preliminary study represents a significant advance in elucidating the molecular pathways of pathologic antibody production in VITT and offers a rare example in human disease of a dangerous small B-cell clone that undergoes rapid clonal expansion and secretion of a harmful monoclonal antibody.^24^ The identification of anti-PF4 proteomic signatures using the workflow herein provides an entry point to analyse these immunological processes at a molecular level; develop novel therapies aimed at removing pathogenic clones (e.g. anti-idiotypes and/or small peptide antibody inhibitors); allow profiling of anti-PF4 clonal evolution invisible to current immunoassays; and enable tracking of individual serum clonotypes by HCDR3 peptide-based quantitative proteomics as reported recently for COVID-19 patients. ^25^

## Data Availability

All data produced in the present study are available upon reasonable request to the authors.

## Acknowledgments

The authors thank the patients for their active participation in this study. J.J.W. is supported by Flinders University DVCR Fellowship and Flinders Health & Medical Research Institute COVID-19 Research Grant.

## Authorship

J.J.W. and T.P.G. developed the study concept, designed the study, compiled the data, performed the analyses, and wrote the paper; B.A. performed the experiments, participated in designing the research, analysing the data and writing of method; T.C. participated in acquisition, analysis and interpretation of data; A.T. participated in study design and interpretation of data; A.C. performed acquisition of data and participated in writing of method; O.Y. recruited and enrolled the patients and performed patient sample collection and diagnostic assays; S.H. participated in developing the study concept and recruiting patient; C.W.T. recruited and enrolled the patients, provided patient information, and participated in designing the research, interpreting the data, and writing the paper; and all authors reviewed the manuscript.

## Data Availability Statement

The data is available upon request through emailing to the corresponding author.

