## Supplementary Information for "Vaccine-induced immune thrombotic thrombocytopenia (VITT) is mediated by a stereotyped clonotypic antibody"

**Content:**

- **Supplementary Methods**
- **References to supplementary methods**
- **Legend to Supplementary Figure 1**
- **Supplementary Figure 1**

### Supplementary Methods

#### Anti-PF4 antibody purification

Antibodies against PF4 were affinity-purified from serum of VITT patients using PF4 protein-coupled MyOne Carboxylic Acid Dynabeads (ThermoFisher). Briefly, the beads were washed twice in 1ml of 15mM MES buffer (pH6) (Sigma) followed by activation with 100μL of 1-Ethyl-3-(3dimethylaminopropyl) carbodiimide (10mg/ml) and incubation on a rotator for 30 min at room temperature. Following activation, the beads were washed with 15mM MES buffer and coated with native PF4 (ChromaTec, Greifswald, Germany) in 15mM MES buffer. Samples were then incubated overnight on a rotator at room temperature. After antigen coating, the beads were washed twice in 0.1% Tween 20 PBS and blocked with PBS 0.1% BSA for 10 min on a rotator. Diluted serum was then added to the beads and mixed on a rotator for 2 h at room temperature. After the mixing, unbound sera were removed, and the beads were washed 4 times with PBS. To elute the antibodies, 100mM glycine elution buffer (pH 11) was added to each sample, vortexed and incubated for 5 min. Eluted anti-PF4 were then transferred to 10kd spin columns (Amicon Ultra) with Milli Q water for buffer exchange and stored at -80ºC until required.

#### Specificity analysis of anti-PF4 antibodies

The activity and specificity of purified anti-PF4 antibodies were determined by testing starting sera (diluted 1:100), eluted anti-PF4 fraction, and unbound fractions (normalized to each starting serum) for reactivity against PF4, SARS-CoV-2 spike S1 and S2 proteins (the Native Antigen Company, UK) by ELISA. In brief, maxisorp nunc immune plates (Thermofisher) were coated with 100 μl of individual PF4, S1 and S2 protein at 4 μg/ml in PBS buffer overnight at 4°C. Plates were blocked with PBS 1%BSA (Sigma-Aldrich, A3059) and subsequently incubated with patient serum (diluted to 1:100) for 2 h at 37 °C. After washing plates four times with PBS 0.05% Tween 20, anti-human IgG (Sigma-A3187) secondary antibodies were added and incubated for 1 h at 37 °C. Plates were then washed six times with PBS 0.05% Tween 20 and phosphatase substrate (Sigma, S0942) was added. Optical density (OD) at 405nm was measured by a plate reader (Spectramax Id5) at 30 minutes of incubation. Blank measurements were subtracted from each sample measurement.

**Mass Spectrometry (MS) sequencing**

Purified anti-PF4 IgG heavy and light-chains were isolated by reduced SDS-PAGE (criterion stain-free TGX gels; Bio-Rad, Hercules, CA, USA). The gel bands were excised and digested with Pierce trypsin protease (ThermoFisher Scientific) and chymotrypsin (Promega), separately.

Peptides were analysed with a Dionex Ultimate 3000 UPLC coupled to a Thermo Fusion Lumos tandem mass spectrometer (Thermo Fisher Scientific, Waltham, Massachusetts, USA). Peptides were applied to a PepMap™ 100 trap cartridge (0.3 x 5 mm, 5 µm C18, Thermo Fischer) and separated on an inhouse 40 cm pulled column created from 75 µm inner diameter fused silica capillary packed with 1.9 µm ReproSil-Pur C18 beads (Dr. Maisch, Ammerbuch, Germany). Solvent A was 0.1% formic acid in water and solvent B was 0.1% formic acid in 80% acetonitrile. For each injection, approximately 1 µg peptides were loaded and separated using a 60-min gradient from 3 to 31.2% B, followed by a 25 min washing and re-equilibration step.

The Fusion Lumos was operated in positive ion mode with a MS1 resolution of 60,000, normalised AGC target of 8e5, scan range of 350-1200m/z and the maximum injection time set to auto for all precursor scans. Data dependent mode was set to a cycle time of 3 seconds with MS/MS performed on the most intense precursors exceeding an intensity threshold of 5e5.

Additional MS/MS settings were a 1.4 m/z quadrupole isolation width, normalised HCD (higher-energy collisional dissociation) energy of 32% and analysis of fragment ions in the orbitrap with a resolution of 15,000, dynamic exclusion was set to 30 seconds, monoisotopic precursor selection (MIPS) was set to Peptide, maximum injection time was set to Dynamic mode, AGC target set to Standard, charge states unknown, +1 or >+7 were excluded, advanced peak determination was toggled on and the EASY-IC internal mass calibration was employed.

Purification of anti-PF4 antibodies from individual patient sera was carried out on at least two independent occasions, and the purified Igs were run for MS at two technical replicates.

**Protein sequence data analysis**

Peptide sequence analysis was performed by de novo sequencing and International ImMunoGeneTics (IMGT) database matching using Peaks studio XPro software (Bioinformatics Solution Inc., Waterloo, ON, Canada) (Figure 1A). Parameters for database searches, data refinement and Ig variable region subfamily assignments were described previously.[1,2] Briefly, a maximum of two missed cleavages, precursor tolerance of < 15 parts per million, product ion tolerance of 0.02 Da, precursor charge state of +2 to +4, fixed modification carbamidomethylation, variable modifications oxidation and deamidation, a maximum of three modifications allowed and non-specific cleavage at one end. High-quality de novo peptides were selected based on sequences having an average local confidence score threshold greater than or equal to 80% and inspected manually to ensure correct assignments. A false discovery rate (FDR) threshold of 1.0% was applied at the peptide level to each data set. The Ig variable region subfamily is assigned from the presence of a unique peptide corresponding to the subfamily.

**Supplementary Figure 1.** Representative annotated MS/MS spectra. The HCDR3 and LCDR3-containing peptides are identified by de novo sequencing in five individual VITT patients (1-5). HCDR3 and LCDR3 regions are highlighted in yellow. Underlined amino acid indicates a post translational modification: M (oxidised methionine), W (oxidised tryptophan), and C (carbamidomethylated cysteine). The sequences, m/z and z of each individual peptides are shown on the top of their annotated MS/MS spectra. Matched b ions are indicated in blue and y ions are in red. m=mass, z=charge.

**VITT 1**

**VHQLGLEDAFDLWGQGT<u>M</u>VTVSSASTK m/z=965.1395 (z=3)**

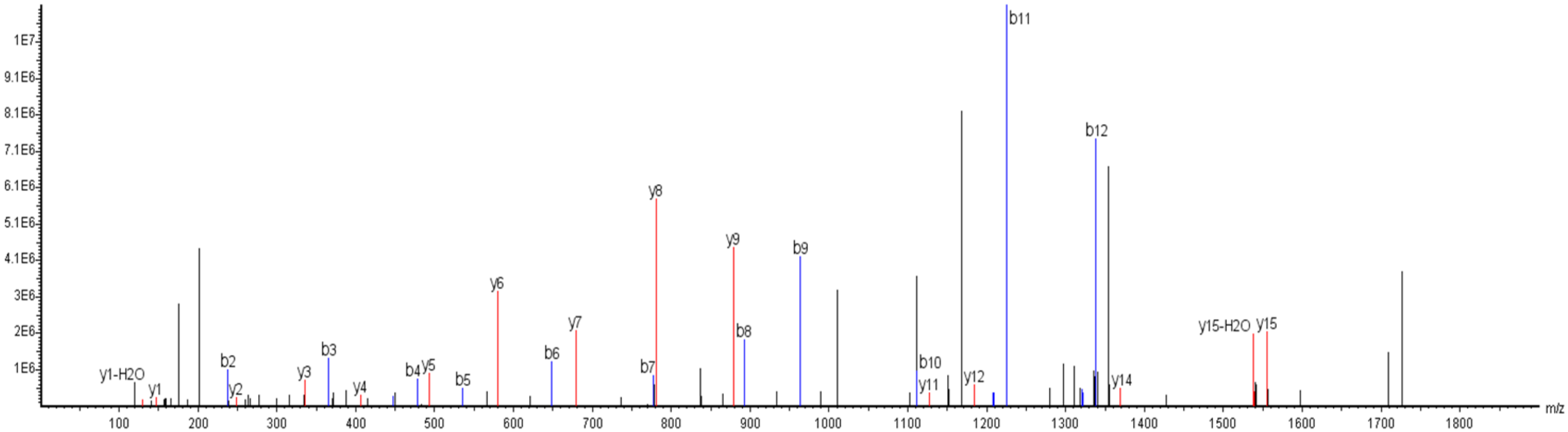


**LHQLGLEDVFDLWGQGTMVTVSSASTK m/z=974.4860 (z=3)**

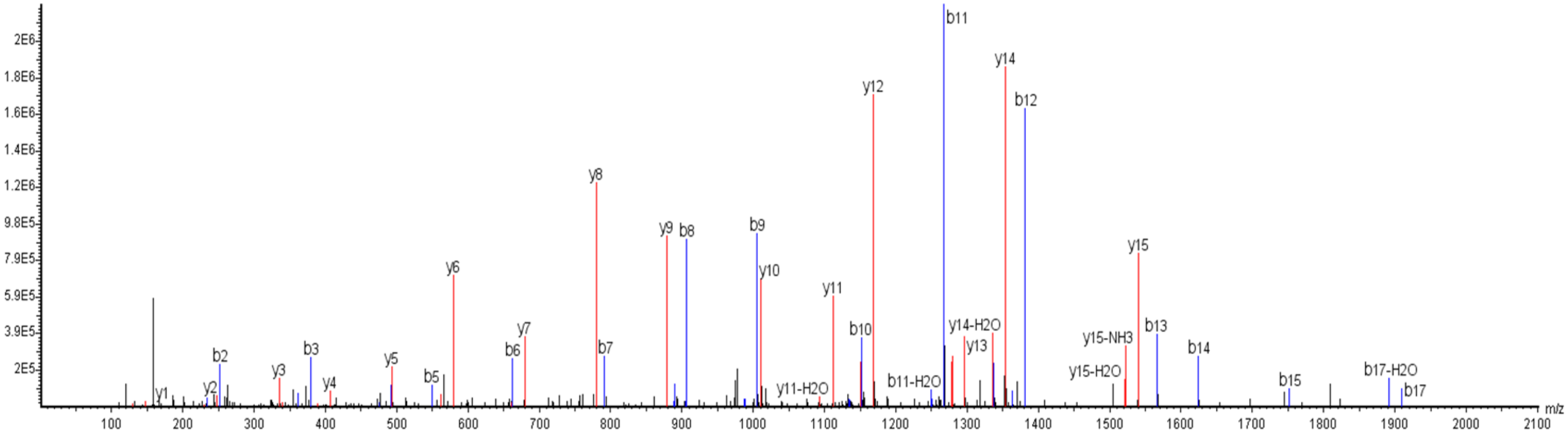

Intensity **VNQLGLEDAFDVWGQGTMVTVSSASTK m/z=947.4679 (z=3)**

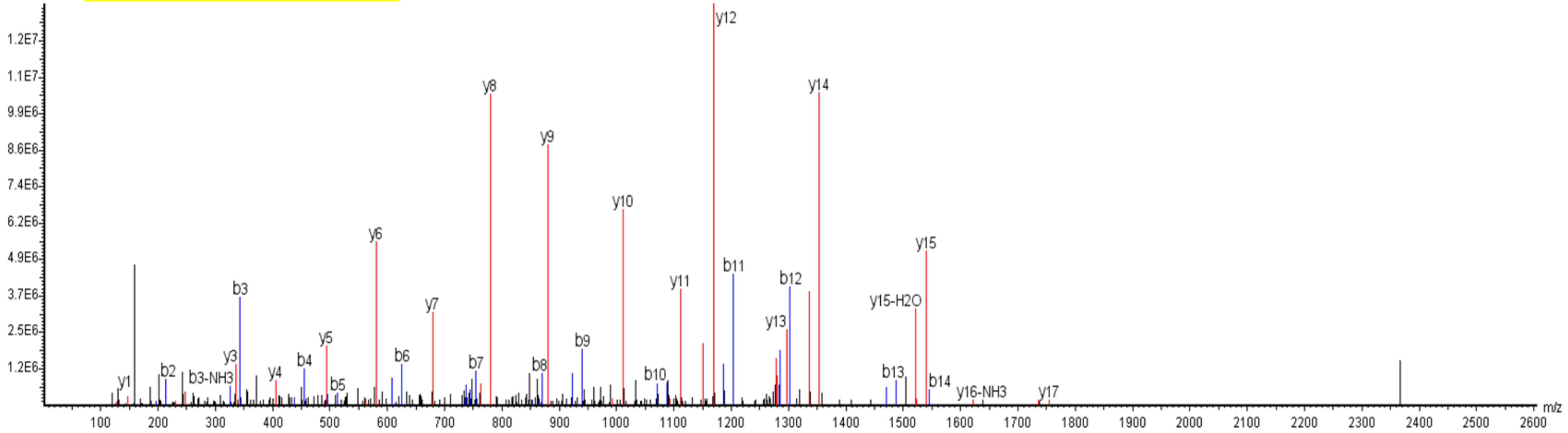


Intensity **VNQLGLEDVFDVWGQGTMVTVSSASTK m/z=962.4755 (z=3)**

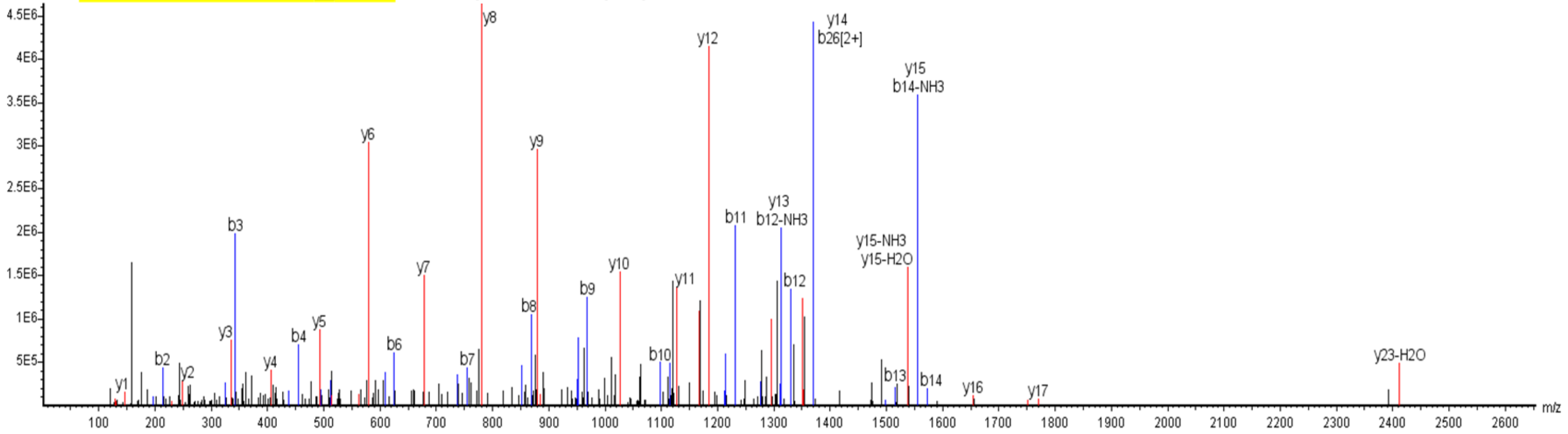

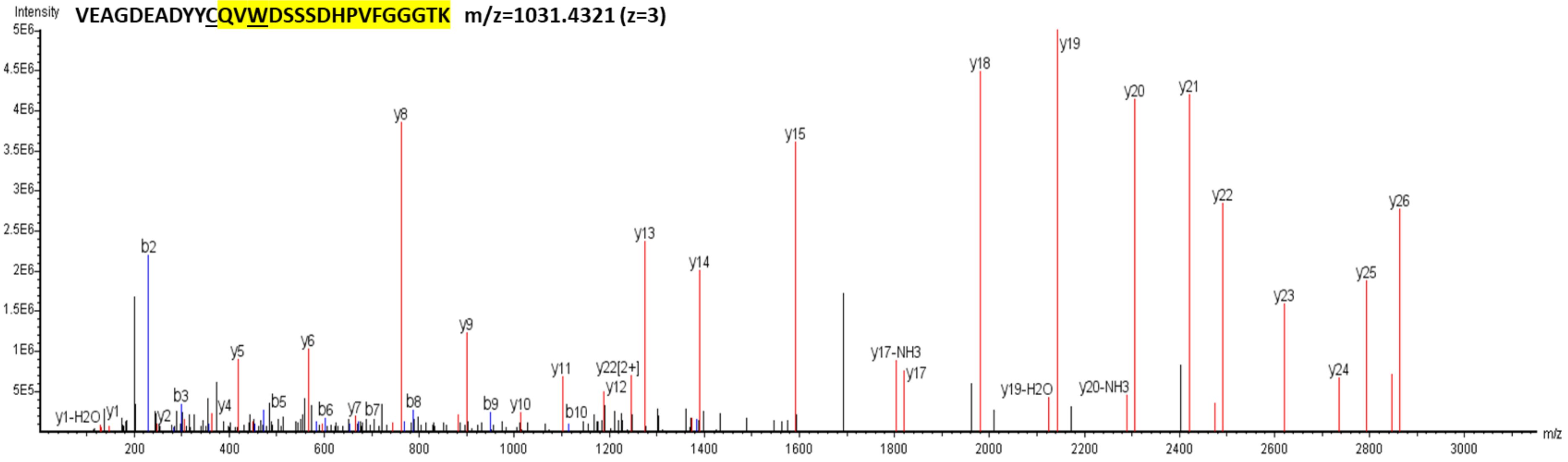
Intensity
VEAGDEADYYCQVWDSSSDHPVFGGGTK m/z=1031.4321 (z=3)
5E6
4.5E6
4E6
3.5E6
3E6
2.5E6
2E6
1.5E6
1E6
5E5
y1-H2O
y1
y2
b2
b3
y4
y5
b5
y6
b6
y7
b7
y8
b8
y9
b9
y10
y11
b10
y22[2+]
y12
y13
y14
y15
y17-NH3
y17
y18
y19-H2O
y19
y20-NH3
y20
y21
y22
y23
y24
y25
y26
m/z
200
400
600
800
1000
1200
1400
1600
1800
2000
2200
2400
2600
2800
3000

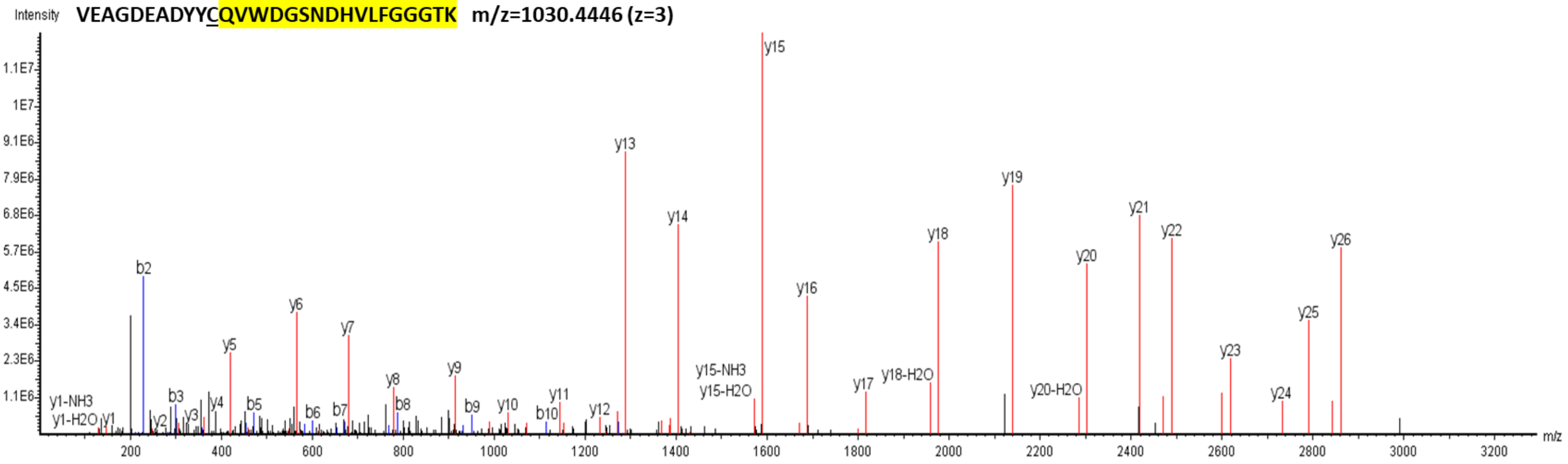
Intensity
VEAGDEADYYCQVWDGSNDHVLFGGGTK m/z=1030.4446 (z=3)
1.1E7
1E7
9.1E6
7.9E6
6.8E6
5.7E6
4.5E6
3.4E6
2.3E6
1.1E6
y1-NH3
y1-H2O
y1
y2
b2
b3
y3
y4
y5
b5
y6
b6
b7
y7
y8
b8
y9
b9
y10
b10
y11
y12
y13
y14
y15-NH3
y15-H2O
y15
y16
y17
y18-H2O
y18
y19
y20-H2O
y20
y21
y22
y23
y24
y25
y26
m/z
200
400
600
800
1000
1200
1400
1600
1800
2000
2200
2400
2600
2800
3000
3200

**VEDGDEADYYCQVWDSSNDHPVFGGGTK m/z=1049.7722 (z=3)**

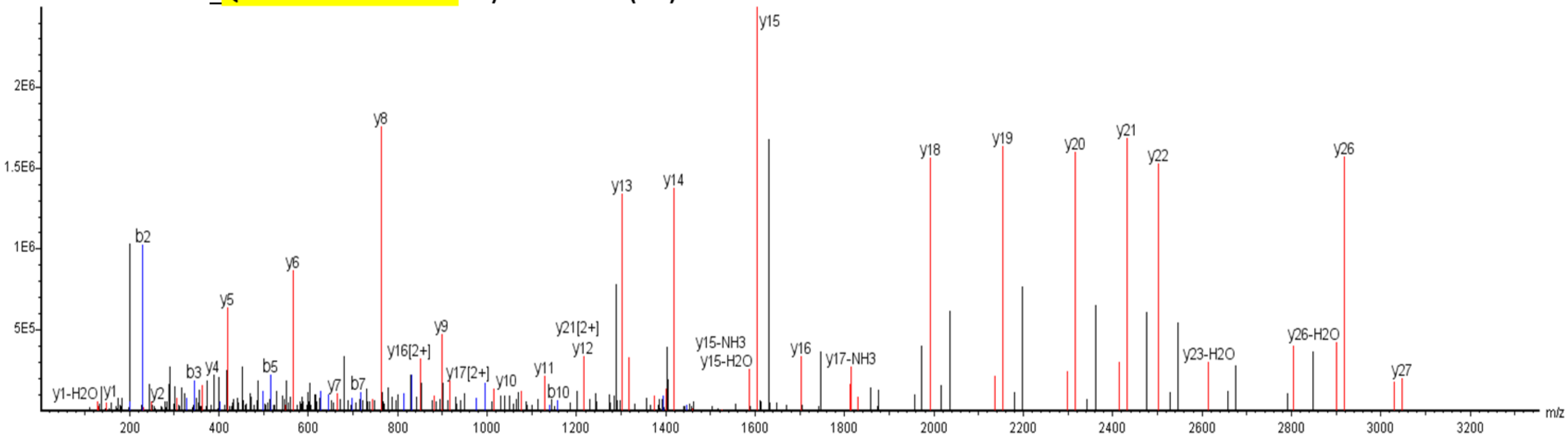


**VEAGDEADYFCQVWDGSSDHPVFGGGTK m/z=1016.0991 (z=3)**

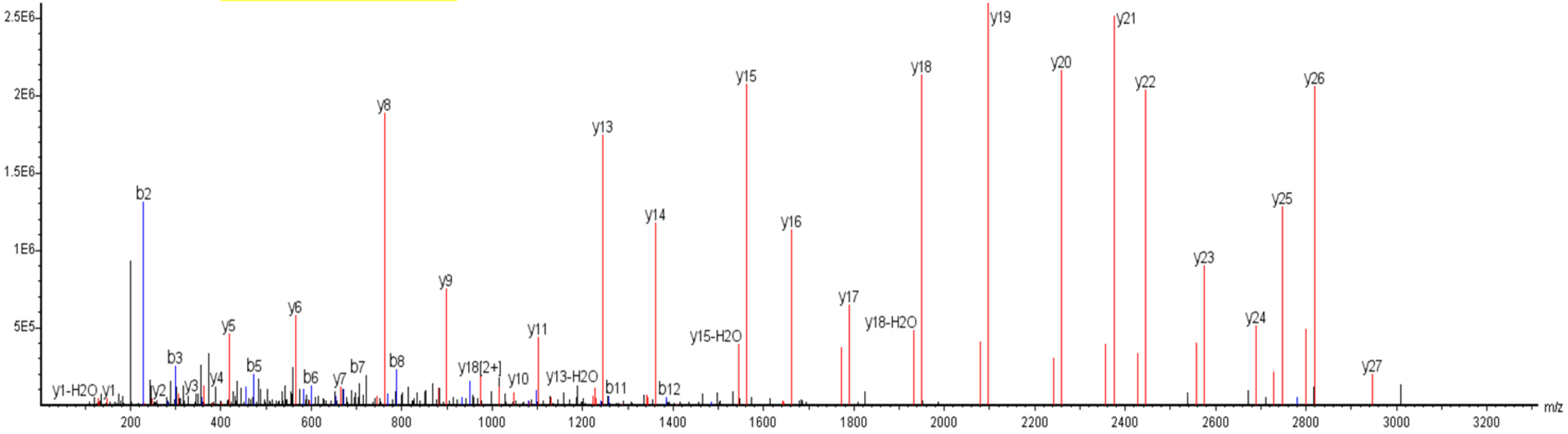

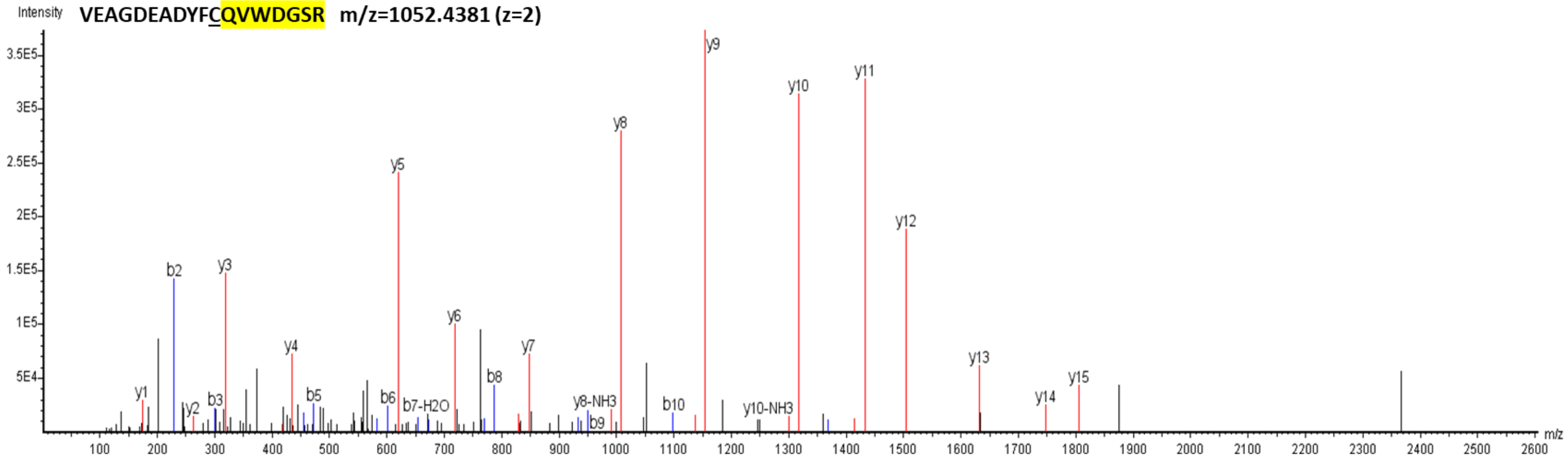
Intensity
VEAGDEADYFCQVWDGSR m/z=1052.4381 (z=2)
3.5E5
3E5
2.5E5
2E5
1.5E5
1E5
5E4
y1 b2 y2 b3 y3 y4 b5 y5 b6 b7-H2O y6 b8 y7 y8-NH3 b9 y8 b10 y9 y10-NH3 y10 y11 y12 y13 y14 y15
100 200 300 400 500 600 700 800 900 1000 1100 1200 1300 1400 1500 1600 1700 1800 1900 2000 2100 2200 2300 2400 2500 2600
m/z

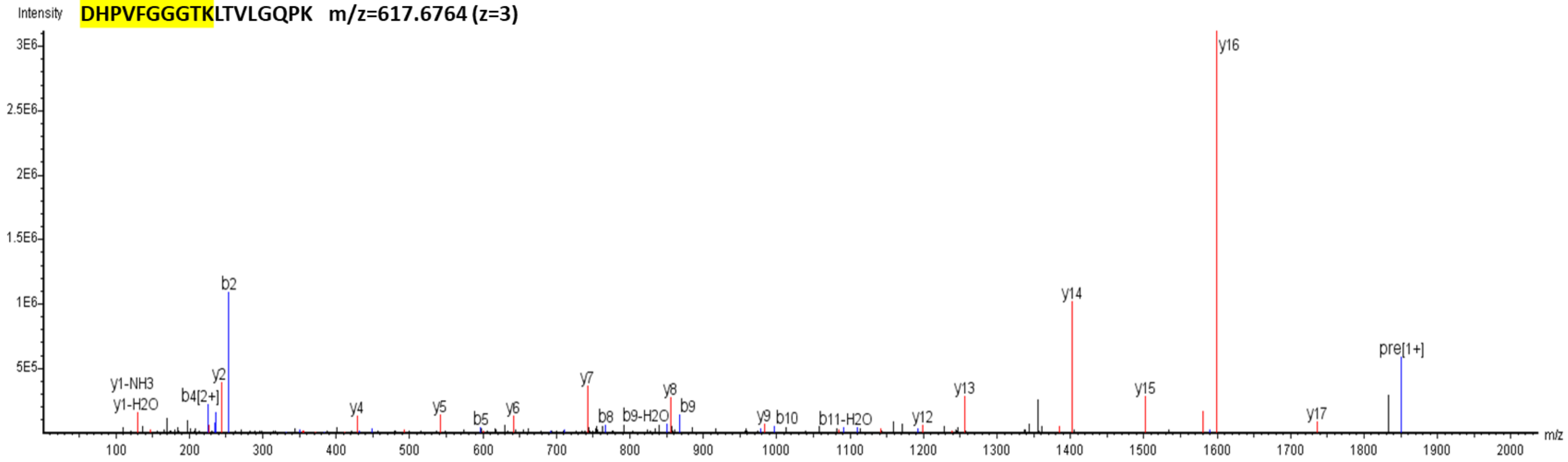
Intensity
DHPVFGGGTKLTVLGQPK m/z=617.6764 (z=3)
3E6
2.5E6
2E6
1.5E6
1E6
5E5
y1-NH3 y1-H2O b4[2+] y2 b2 y4 y5 b5 y6 y7 b8 b9-H2O y8 b9 y9 b10 b11-H2O y12 y13 y14 y15 y16 y17 pre[1+]
100 200 300 400 500 600 700 800 900 1000 1100 1200 1300 1400 1500 1600 1700 1800 1900 2000
m/z

**VITT 2**

**VVGLEDAFDLWGQGTMLTVSSASTK m/z=871.4385 (z=3)**

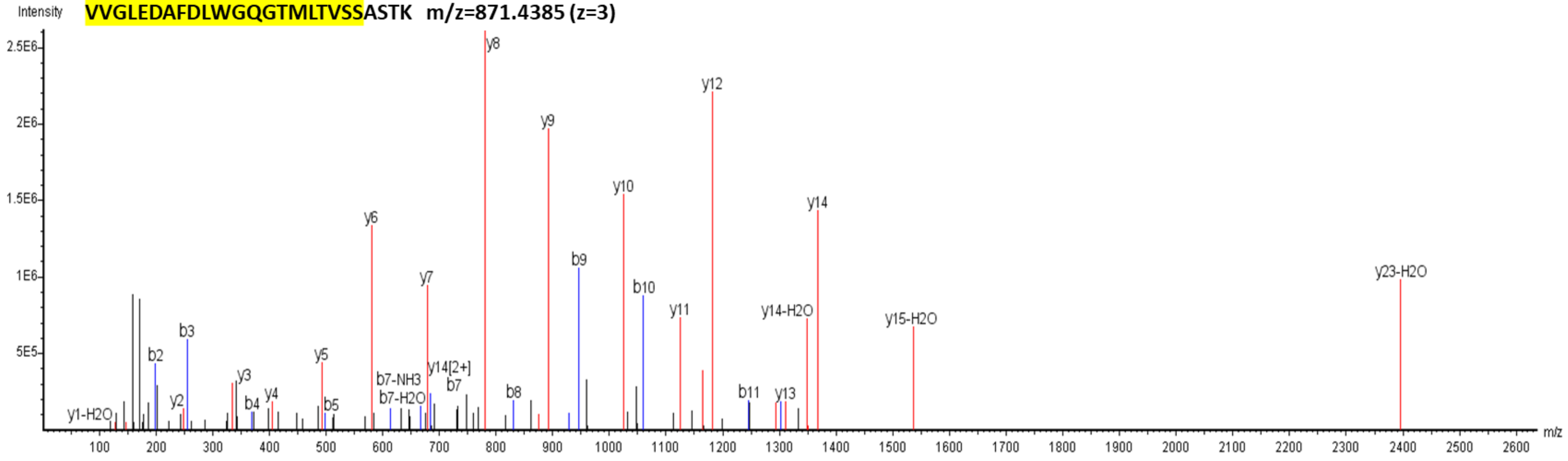


**ERVVGLEDAFDFR m/z=776.8851 (z=2)**

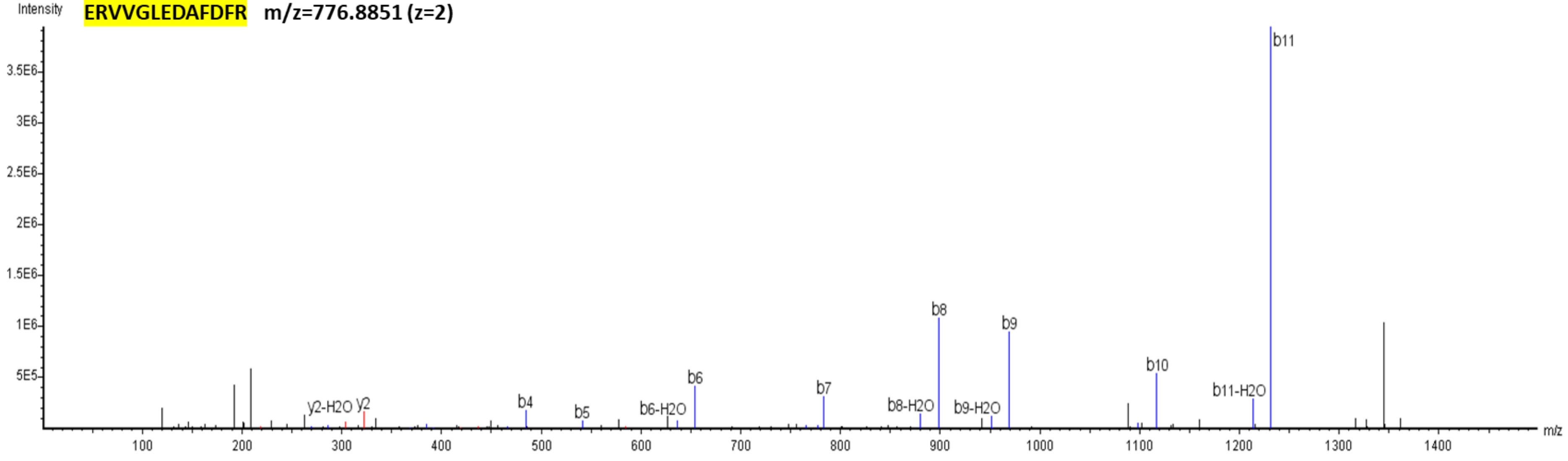

**VVGLEDAFDFRGQGTMLTVSSASTK m/z=878.1004 (z=3)**

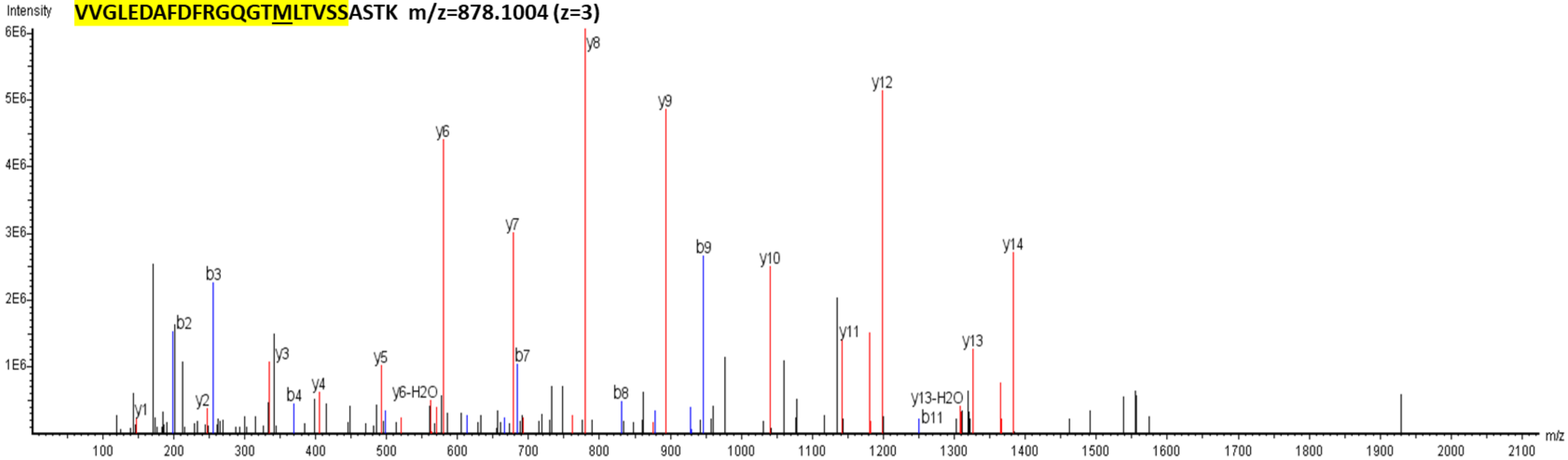


**VEAGDEADYYCQMWDGSSDHPLFGGGTK m/z=1031.4277 (z=3)**

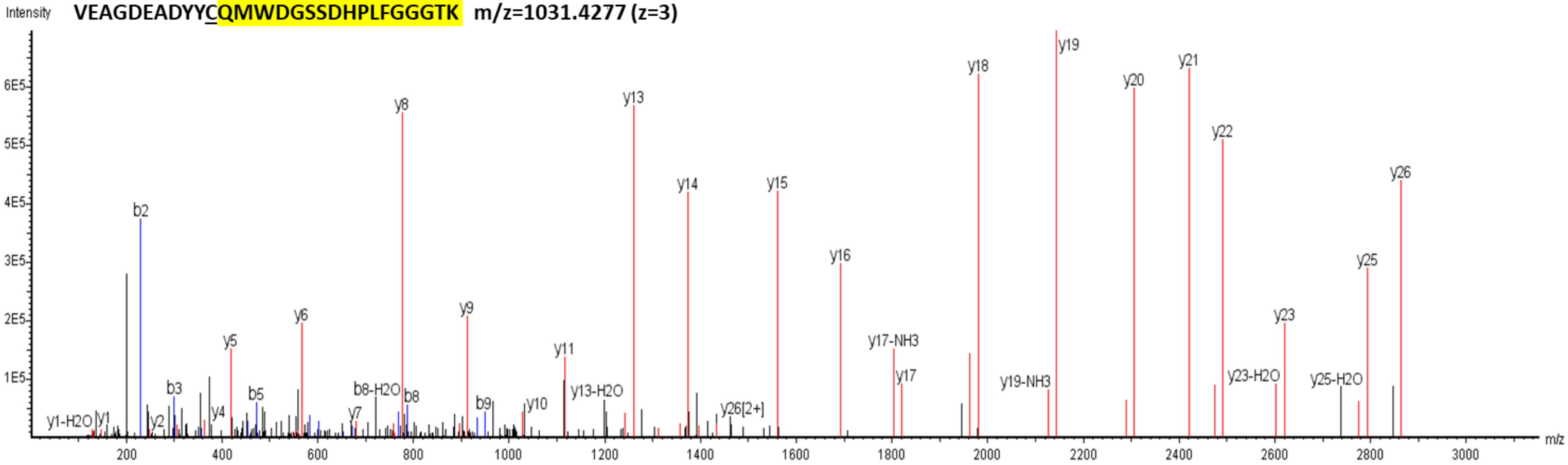

**VITT 3**

**LFQLNLEDAFDLWGQGTMVTVSSASTK   m/z=992.1530 (z=3)**

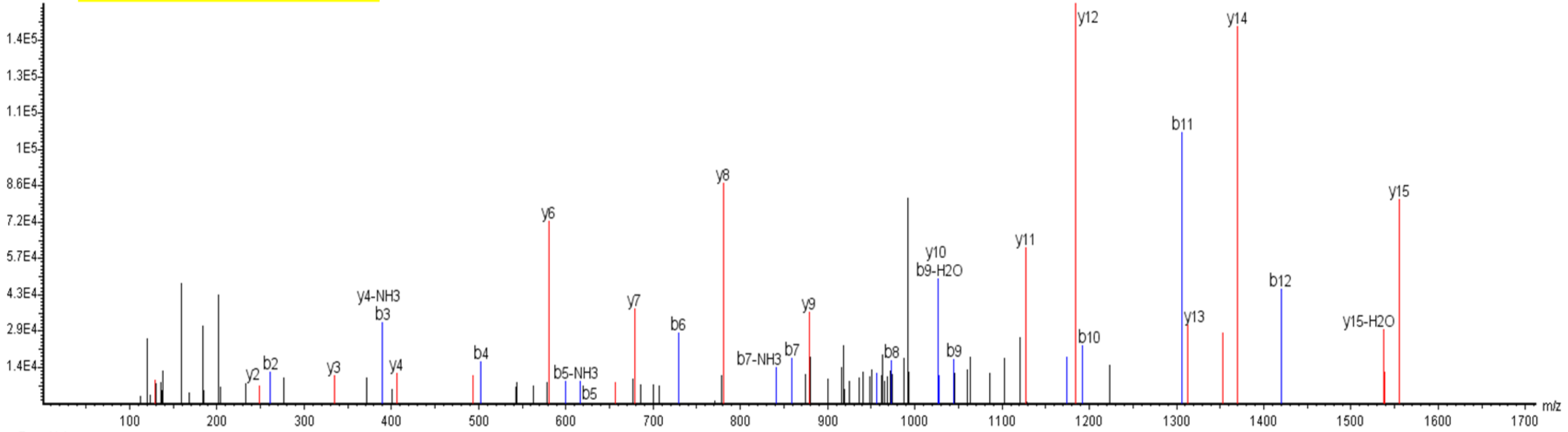


**LFQLNLEDAFDFRGQGTMVTVSSASTK   m/z=993.4881 (z=3)**

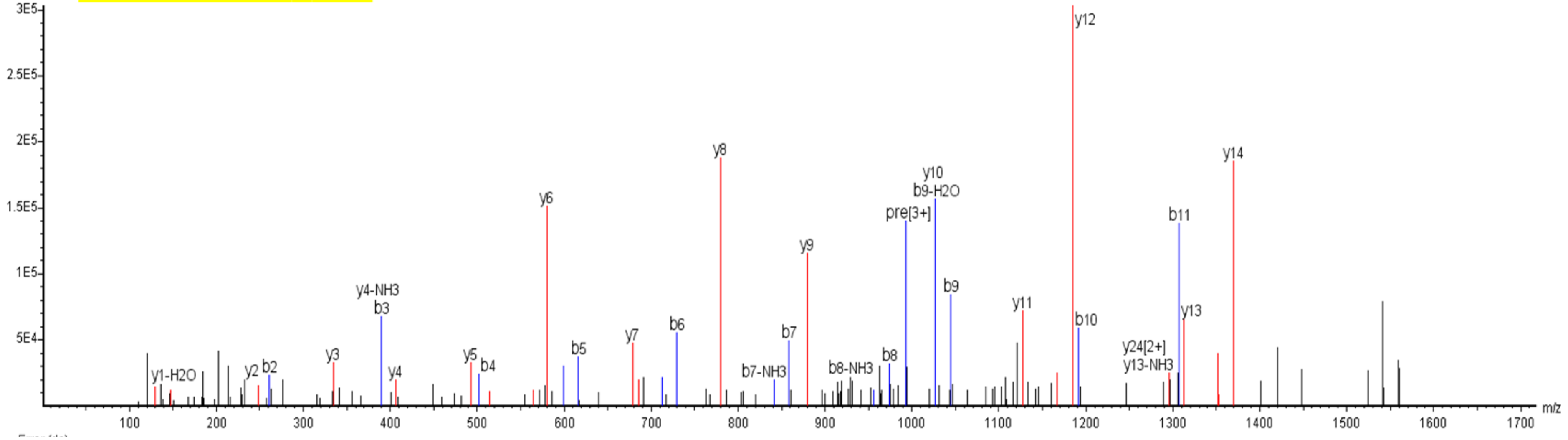

**VEAGDEADYY<u>C</u>QVWDGSNDHVLFGGGTK m/z=1030.4456 (z=3)**

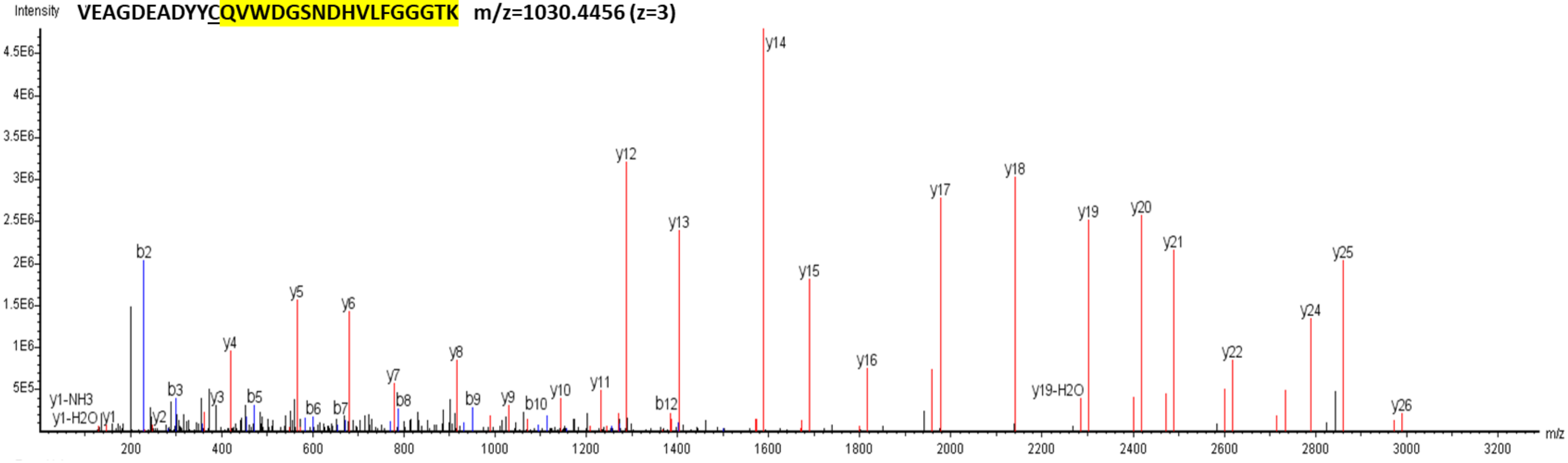

**VITT 4**

Intensity **LFQLGLEDTFDLWGQGTLVTLSSPSTK m/z=990.5077 (z=3)**

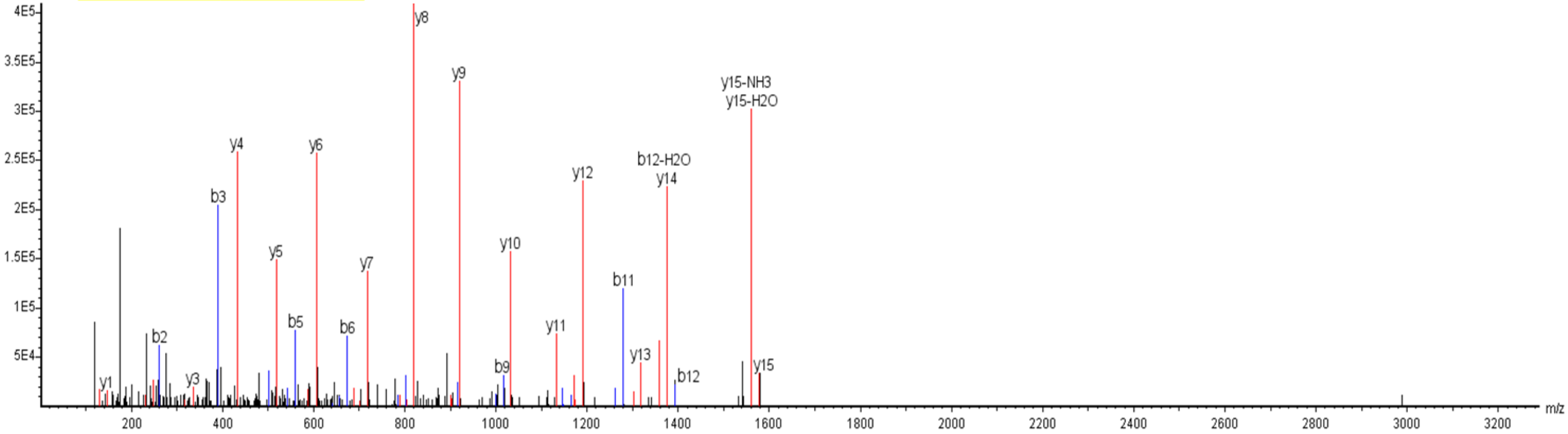


Intensity **VFQLGLEDTFDLWGQGTLVTLSSPSTK m/z=980.5062 (z=3)**

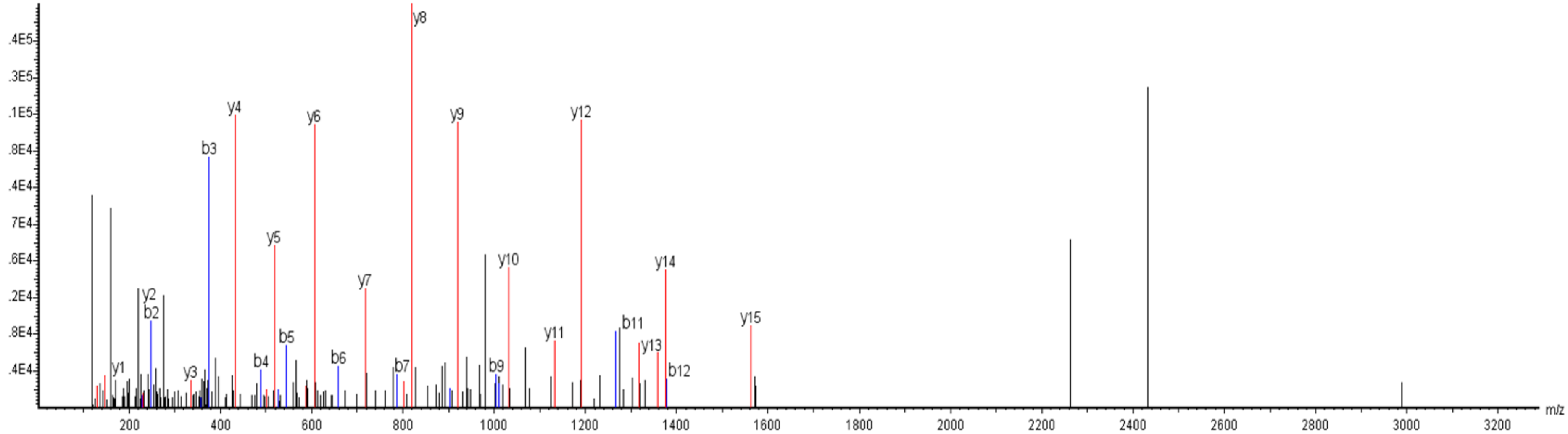

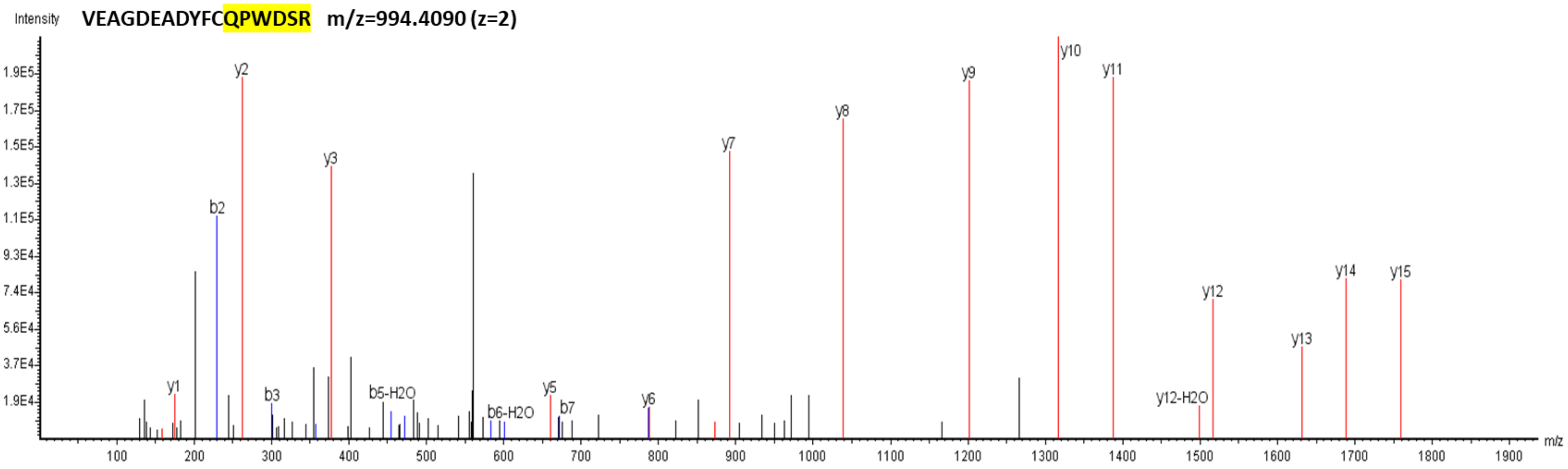
Intensity
VEAGDEADYFCQPWDSR m/z=994.4090 (z=2)
1.9E5
1.7E5
1.5E5
1.3E5
1.1E5
9.3E4
7.4E4
5.6E4
3.7E4
1.9E4
y1
b2
y2
b3
y3
b5-H2O
b6-H2O
y5
b7
y6
y7
y8
y9
y10
y11
y12-H2O
y12
y13
y14
y15
m/z
100
200
300
400
500
600
700
800
900
1000
1100
1200
1300
1400
1500
1600
1700
1800
1900

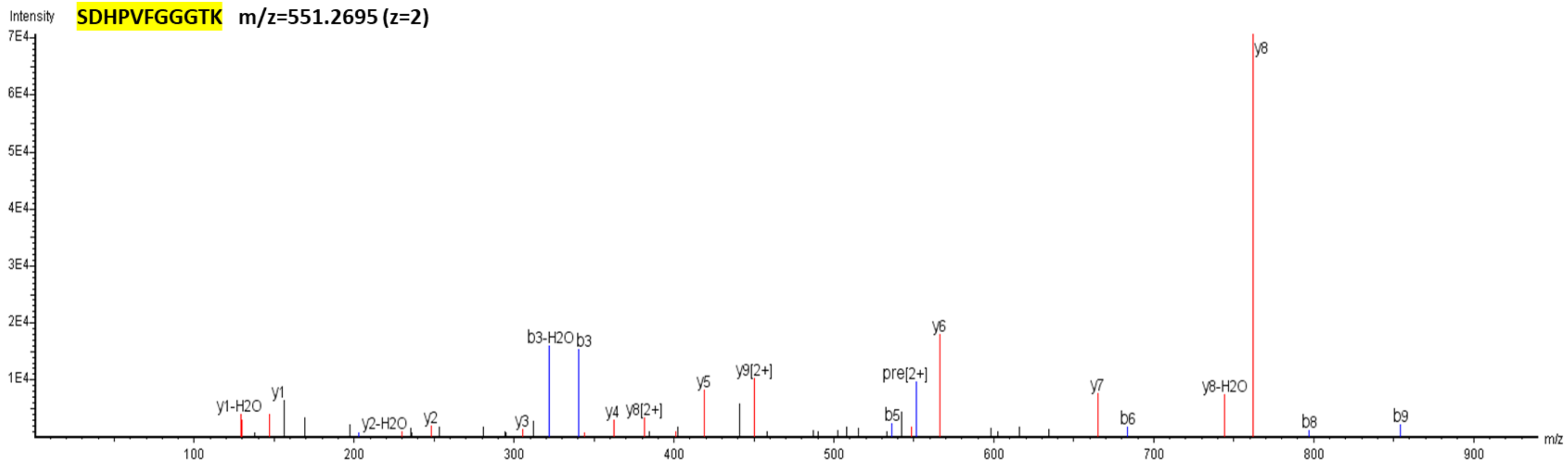
Intensity
SDHPVFGGGTK m/z=551.2695 (z=2)
7E4
6E4
5E4
4E4
3E4
2E4
1E4
y1-H2O
y1
y2-H2O
y2
y3
b3-H2O
b3
y4
y8[2+]
y5
y9[2+]
b5
pre[2+]
y6
y7
b6
y8-H2O
y8
b8
b9
m/z
100
200
300
400
500
600
700
800
900

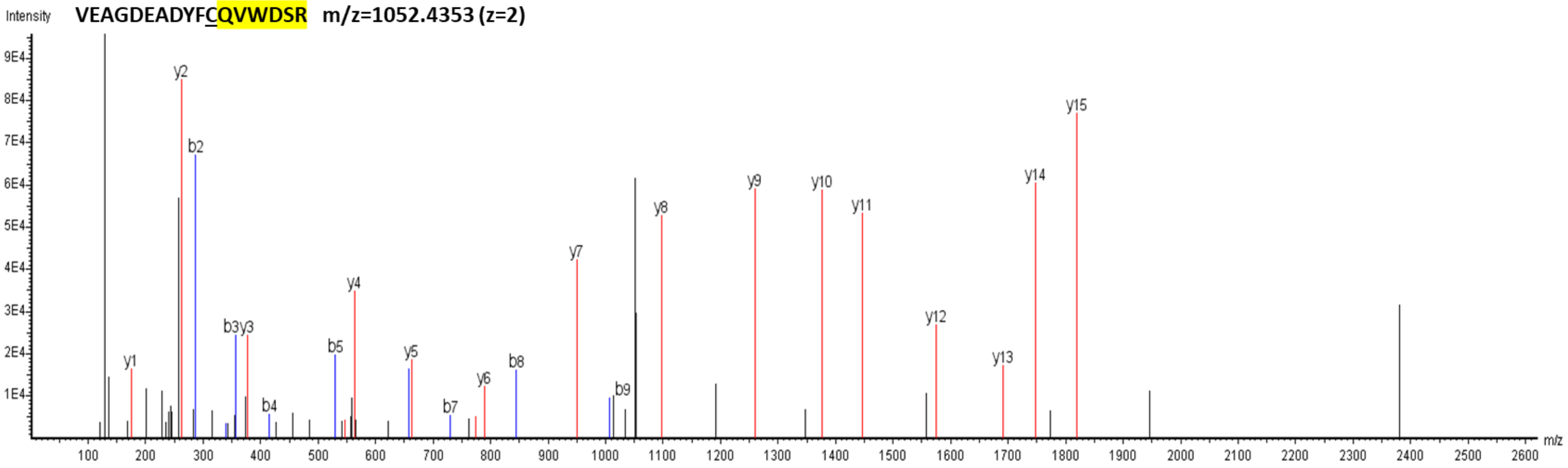

Intensity
VEAGDEADYFCQVWDSR m/z=1052.4353 (z=2)
9E4
8E4
7E4
6E4
5E4
4E4
3E4
2E4
1E4
y1
y2
b2
b3y3
b4
b5
y4
y5
b7
y6
b8
y7
b9
y8
y9
y10
y11
y12
y13
y14
y15
100
200
300
400
500
600
700
800
900
1000
1100
1200
1300
1400
1500
1600
1700
1800
1900
2000
2100
2200
2300
2400
2500
2600
m/z

**VITT 5**

**AHVLGLEDVFDLWGQGTMVTVSSASTK m/z=955.8082 (z=3)**

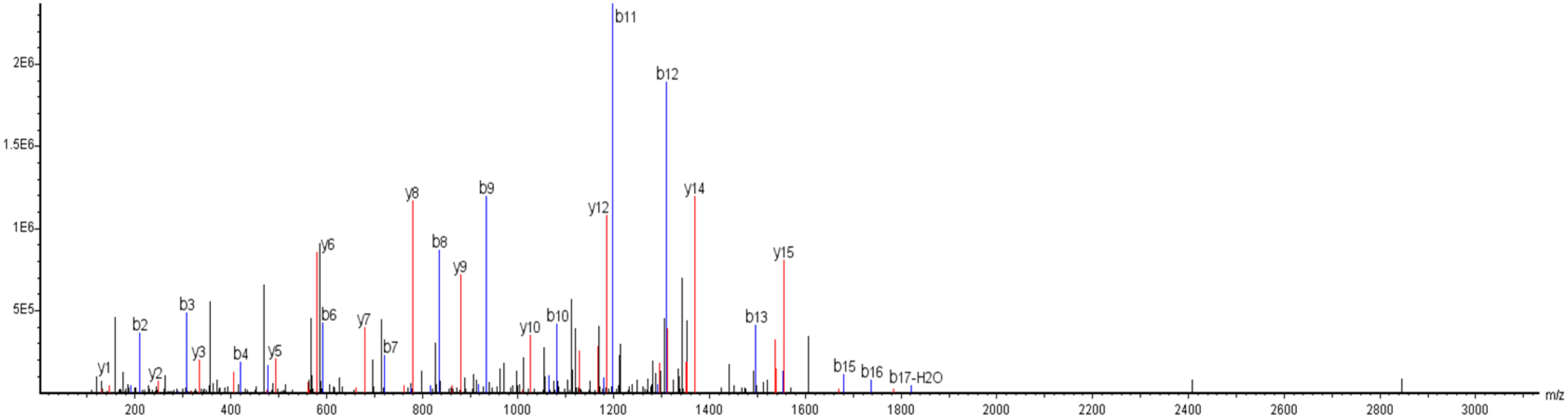


**AHVLGLEDAFDLWGQGTMVTVSSASTK m/z=951.7988 (z=3)**

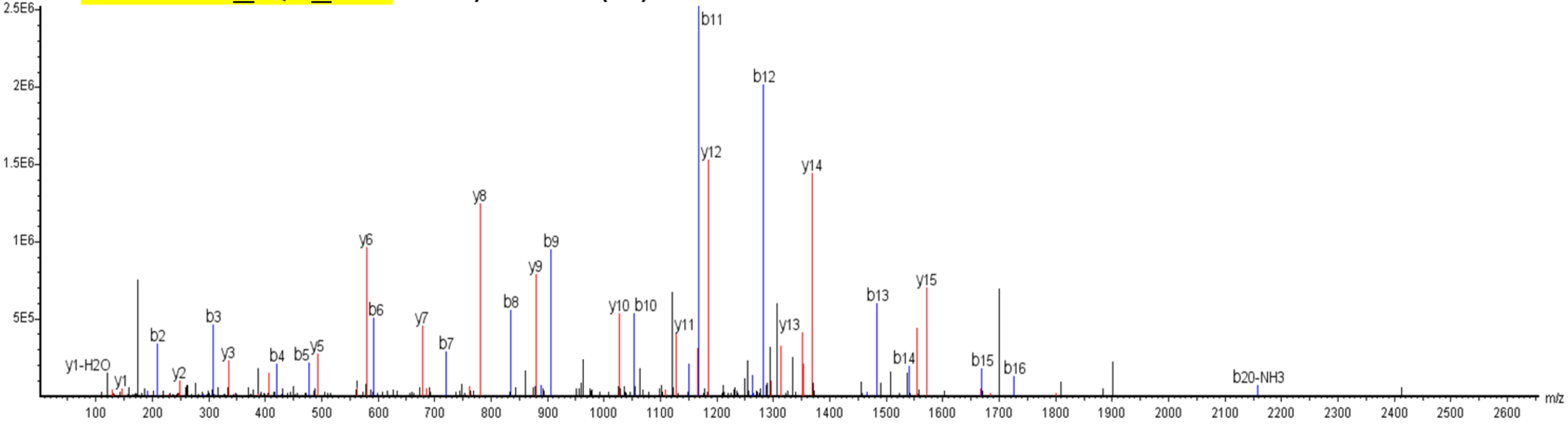

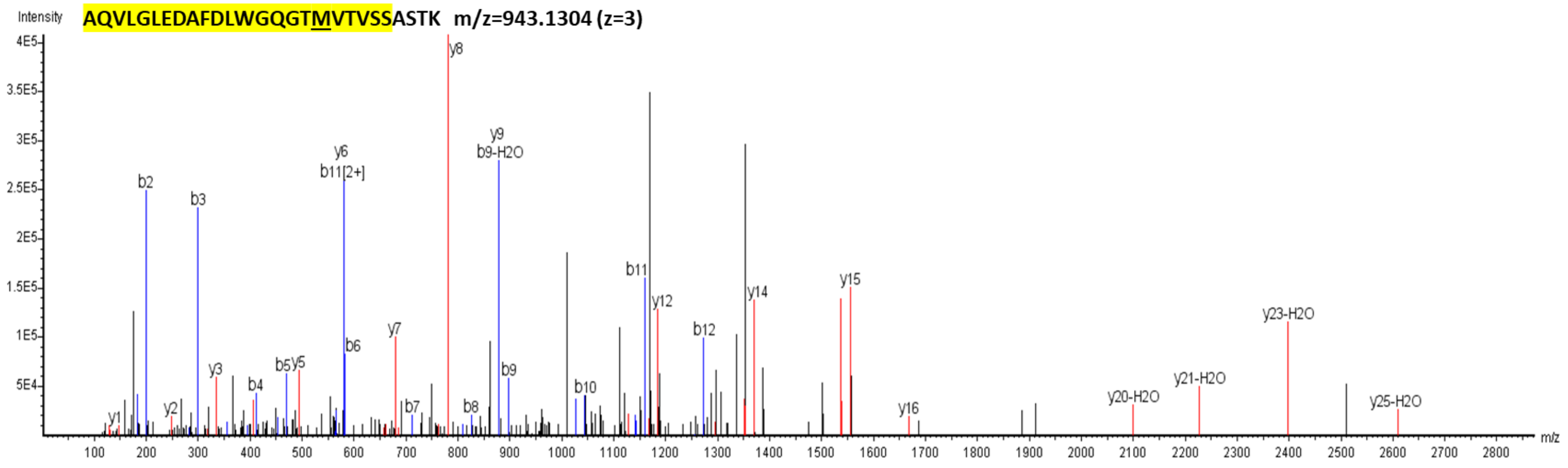

AQVLGLEDAFDLWGQGTMVTVSSASTK m/z=943.1304 (z=3)
Intensity
m/z
y8
y9
b9-H2O
y6
b11[2+]
b2
b3
b11
y12
y14
y15
b12
y7
b6
b5
y5
y3
b4
b9
b10
y1
y2
b7
b8
y16
y20-H2O
y21-H2O
y23-H2O
y25-H2O


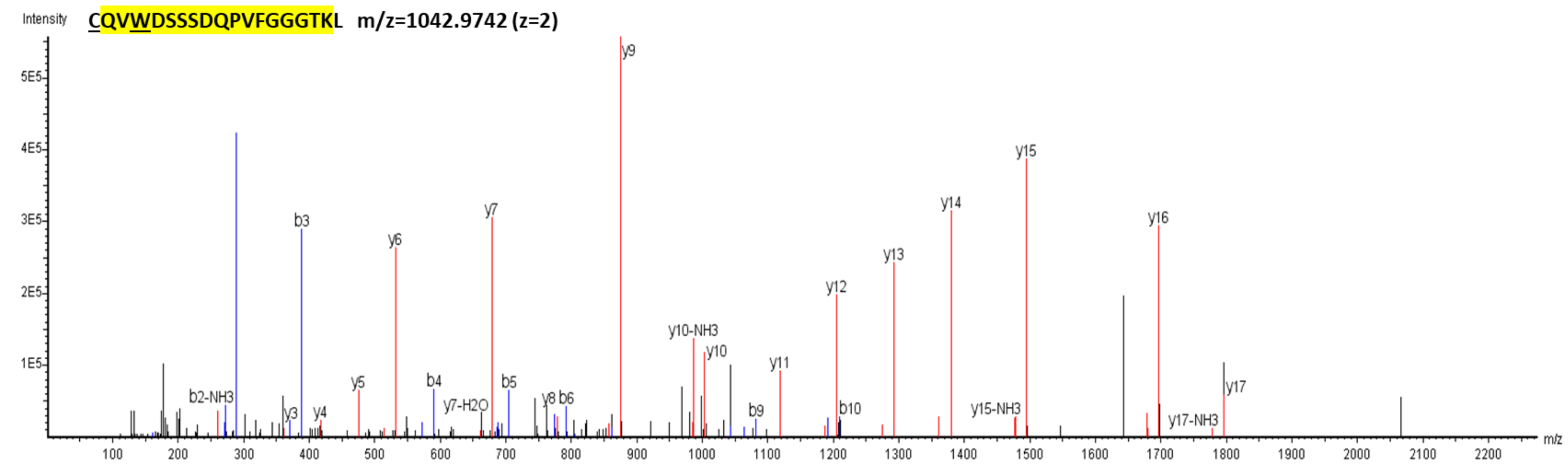

CQVWDSSSDQPVFGGGTKL m/z=1042.9742 (z=2)
Intensity
m/z
y9
y15
y14
y16
y7
b3
y6
y13
y12
y10-NH3
y10
y11
b2-NH3
y5
b4
b5
y3
y4
y7-H2O
y8
b6
b9
b10
y15-NH3
y17
y17-NH3

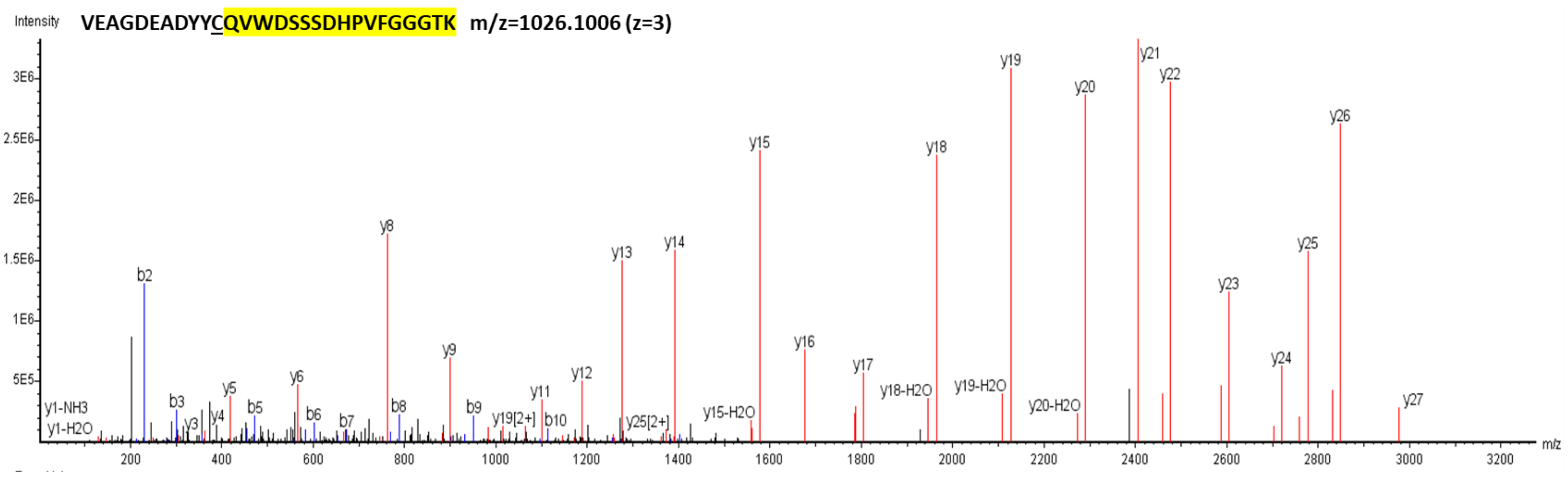

VEAGDEADYYCQVWDSSSDHPVFGGGTK m/z=1026.1006 (z=3)
Intensity
3E6
2.5E6
2E6
1.5E6
1E6
5E5
y1-NH3
y1-H2O
b2
b3
y3
y4
y5
b5
y6
b6
b7
y8
b8
y9
b9
y19[2+]
y11
b10
y12
y13
y25[2+]
y14
y15-H2O
y15
y16
y17
y18-H2O
y18
y19-H2O
y19
y20-H2O
y20
y21
y22
y23
y24
y25
y26
y27
200
400
600
800
1000
1200
1400
1600
1800
2000
2200
2400
2600
2800
3000
3200
m/z
